# Doxycycline and Hydroxychloroquine as Treatment for High-Risk COVID-19 Patients: Experience from Case Series of 54 Patients in Long-Term Care Facilities

**DOI:** 10.1101/2020.05.18.20066902

**Authors:** Imtiaz Ahmad, Mohammud Alam, Ryan Saadi, Saborny Mahmud, Emily Saadi

## Abstract

**Importance:** Patients in long-term care facilities (LTCF) are at a high-risk of contracting COVID-19 due to advanced age and multiple comorbidities. Without effective treatments, outbreaks in such facilities will become commonplace and will result in severe morbidity and mortality. The effectiveness of doxycycline (DOXY) and hydroxychloroquine (HCQ) combination therapy in high risk COVID-19 patients in longterm care facilities is not yet understood.

**Objective:** The goal of this analysis is to describe outcomes after use of DOXY-HCQ combination in high-risk COVID-19 patients in LTCF.

**Design:** Case-series analysis.

**Setting:** Three (3) LTCFs in New York.

**Participants:** From March 19 to March 30, 2020, fifty-four (54) patients, residents of three (3) LTCFs in New York and diagnosed (confirmed or presumed) with COVID-19, were included in this analysis.

**Exposure:** All patients who were diagnosed (confirmed or presumed) with COVID-19 received DOXY-HCQ combination therapy along with standard of care.

**Main Outcomes and Measures:** Patients characteristics, clinical recovery, radiological improvements, medication side-effects, hospital transfer, and death were assessed as outcome measures.

**Results:** A series of fifty-four (54) high-risk patients, who developed a sudden onset of fever, cough, and shortness of breath (SOB) and were diagnosed or presumed to have COVID-19, were started with a combination of DOXY-HCQ and 85% (n=46) patients showed clinical recovery defined as: resolution of fever and SOB, or a return to baseline setting if patients are ventilator-dependent. A total of 11% (n=6) patients were transferred to acute care hospitals due to clinical deterioration and 6% (n=3) patients died in the facilities. Naive Indirect Comparison suggests these data were significantly better outcomes than the data reported in MMWR (reported on March 26, 2020) from a long-term care facility in King County, Washington where 57% patients were hospitalized, and 22% patients died.

**Conclusion:** The clinical experience of this case series indicates DOXY-HCQ treatment in high-risk COVID-19 patients is associated with a reduction in clinical recovery, decreased transfer to hospital and decreased mortality were observed after treatment with DOXY-HCQ.

## Introduction

The COVID-19 pandemic is straining the U.S. healthcare system due to increased hospitalization, ICU admissions, and mortality. As of May 18, 2020, a total of 351,371 patients and 1,508,308 patients were afflicted with COVID-19 in New York state and the United States, respectively^1^. In addition, 75,870 patients were hospitalized, and 28,339 patients died in New York alone^1^. Thus, New York has become the epicenter of the world for the COVID-19 pandemic at the time of this publication^2^.

The COVID-19 virus affects the upper and lower respiratory tract, leading to acute respiratory failure. Consequently, pro-inflammatory cytokines are released such as tumor necrosis factor (TNF), interferon-alpha (IFNa), IL-1β, and IL-6, all of which are mediators of lung inflammation and fever^3^. Therefore, symptoms of a COVID-19 infection include fever, cough, and shortness of breath (SOB). In addition, the virus is associated with acute cardiac injury in 19.7% of hospitalized patients, manifesting as an ejection fraction decline and an elevation in troponin I levels^4^.

Recent small studies in France and China demonstrated the limited efficacy of hydroxychloroquine (HCQ) alone or in combination with azithromycin (AZ)^5,6,7^. However, concerns have been raised about the use of the AZ-HCQ combination due to higher arrhythmogenic potential in high-risk patients with pre-existing cardiac comorbidities and/or acute cardiac injury due to COVID-19. According to the Food and Drug Administration (FDA) labeling, side-effects of AZ include “prolonged cardiac repolarization and QT interval, imparting a risk of developing cardiac arrhythmia and torsades de pointes”^8^. Additionally, QTc prolongation was found in a dose-dependent manner when chloroquine, an HCQ analogue, was used alone or in combination with AZ^8^.

Doxycycline (DOXY) is considered to be an anti-inflammatory, immunomodulatory, and cardioprotective drug ^19^. Both *in-vitro and in-vivo* studies demonstrated that doxycycline directly modulates the expression of inflammatory mediators involved in COVID-19 pathophysiology (IL-8, IL-1β^9^, TNF^10^, MMP^11^, and IL-6^12^). In addition, Kalish et al showed a novel immunosuppressive mechanism for minocycline (a DOXY analogue), as well as its possible additive anti-inflammatory effect when combined with chloroquine or HCQ^20^. No cardiac side-effects are noted per the FDA labeling of DOXY^13^. In addition, during reperfusion after myocardial injury, an acute release of matrix metalloproteinases-2 (MMP-2) is noted^14^, and MMP inhibition after myocardial infarction yields preservation of left ventricular function^15^. DOXY reduces the adverse LV remodeling in patients with acute ST-segment elevation myocardial infarct (STEMI) and LV dysfunction, possibly via MMP inhibition pathway^16^. These potential cardioprotective effects may have an additional implication in the COVID-19 patients with acute myocardial injury.

Due to these scientific findings, we have undertaken a thorough review of this case series of high-risk COVID-19 patients in nursing homes, who were treated with a combination of DOXY-HCQ.

## Methods

From March 19 to March 30, 2020, we analyzed the clinical outcomes of fifty-four (54) high-risk patients who developed a sudden onset of fever, cough, and SOB, were diagnosed or presumed to have COVID-19, and were treated with DOXY (100 mg PO BID for 7 days) and HCQ (two regimens: i) 200 mg PO TID for 7 days or ii) 400 mg PO BID one day, then 400 mg daily for 6 days). The patients were residents of three (3) long term care facilities (LTCF) in New York. Patients’ characteristics, clinical recovery, radiological improvements, medication side-effects, hospital transfer, and death were assessed as outcome measures in this population. Consent was obtained from all patients and/or families before starting Doxycycline and Hydroxychloroquine to treat presumed and confirmed COVID-19 infection and approved by the oversight medical boards. The retrospective review was approved by Corporate Clinical Services.

High-risk patients were defined as patients who had at least one comorbidity, such as hypertension, diabetes, coronary artery disease (CAD), congestive heart failure (CHF), chronic obstructive pulmonary disorder (COPD), and cerebrovascular accident (CVA). Clinical recovery was defined as a resolution of fever and SOB, or a return to baseline settings for ventilator-dependent patients. A return to baseline ventilator settings was defined as a return of oxygen saturation and mode of ventilator to a baseline level. Chest X-rays were ordered at the onset of symptoms. Follow-up chest r-rays were ordered if clinically indicated.

## Results

In this series of fifty-four (54) patients, the median age was 67 yrs. (range 22-97 yrs.). **Table 1** summarizes the characteristics and outcomes of all COVID-19 patients treated with DOXY-HCQ combination in LTCFs in New York. 96% (n=52) tested positive for COVID-19 and 4% (n=2) were presumed to have the viral infection due to clinical features. 85% (n=46) of patients showed clinical recovery, defined by resolution of fever and SOB, or a return to baseline ventilator setting. 6% (n=3) of patients died and 11% (n=6) of patients were transferred to acute care hospitals due to clinical deterioration.

**Table 1:** Characteristics and Outcome of COVID-19 Patients Treated with DOXY-HCQ Combination in Three (3) Long Term Care Facilities.

| Characteristics | LTCF Resident (n=54) | % | Reference Population (LTACH,WA) |
| --- | --- | --- | --- |
| Median Age, yrs. (range) | 67 (22-97) |  |  |
| **Sex** |  |  |  |
| Male | 33 | 61% |  |
| Female | 21 | 39% |  |
| **Comorbidities** |  |  |  |
| Hypertension | 49 | 91% |  |
| Diabetes | 21 | 39% |  |
| CAD | 31 | 57% |  |
| CHF | 11 | 20% |  |
| CVA | 20 | 37% |  |
| COPD | 21 | 39% |  |
| Vent dependent*** | 17 | 31% |  |
| **Test Status** |  |  |  |
| COVID-19 Positive | 52 | 96% |  |
| COVID-19 Presumed | 2 | 4% |  |
| **Outcome** |  |  |  |
| Clinically recovered** | 46 | 85% |  |
| Transferred to hospital | 6 | 11% | 57% |
| Death | 3 | 6% | 22% |
| **Radiology** |  |  |  |
| Abnormal CXR c/w Pneumonia | 36 | 67% |  |
| Chest x-ray improved | 6 | 11% |  |
| Chest x-ray not improved<br>(possible lag time) | 16 | 30% |  |
| Unavailable* | 32 | 59% |  |
| **Medication side-effect** |  |  |  |
| Seizure | 1 | 2% |  |
| None | 50 | 93% |  |
| Unknown**** | 3 | 6% |  |

\* Chest x-ray either not ordered, or not required or pending
\*\* Recovery defined by resolution of fever and shortness of breath, or return to baseline vent setting
\*\*\* 65% (n =11) patients recovered and vent setting is returned to baseline
\*\*\*\* Not available as patients were transferred to hospital
! Dropouts means patients who did not complete a full course of DOXY-HCQ therapy due to transfer to hospital, death or side-effects.

All patients became and remained afebrile within 0-5 days and SOB resolved or returned to baseline with 0-6 days of DOXY-HCQ initiation. 11% of patients (n=6) showed an improvement in chest X-rays. 30% of patients (n=16) did not show chest X-ray improvement. 93% (n=50) did not display any side-effects of DOXY-HCQ. 2% (n=1) had a seizure and HCQ was immediately terminated.

There were 9 patients who did not complete the 7-day course of DOXY-HCQ due to hospital transfers, death, or side-effects **(Table 2)**. Excluding these patients, a total of 45 patients completed the DOXY-HCQ combination therapy; all 45 patients clinically recovered **(Table 3)**.

**Table 2:** Characteristics COVID-19 Patients Who Did Not Complete DOXY-HCQ Combination Therapy in Three (3) Long Term Care Facilities

|  | **Transferred to hospital** | **Death** | **Details** |
| --- | --- | --- | --- |
| 1 | Yes | No | Transferred to hospital with septic shock, patient returned to nursing facility |
| 2 | Yes | No | Transferred to hospital with septic shock, patient returned to nursing facility |
| 3 | Yes | Unknown* | Transferred to hospital with respiratory failure |
| 4 | No | No | HCQ discontinued due to seizure, patient continued with DOXY. Stable |
| 5 | No | Yes | Died due to respiratory failure at nursing home |
| 6 | Yes | Unknown* | Patient coded, was resuscitated and transferred to hospital |
| 7 | Yes | Yes | Transferred to hospital with respiratory failure and later died in hospital |
| 8 | Yes | No | Transferred to hospital with respiratory failure, patient is in ICU |
| 9 | No | Yes | Died due to septic shock in nursing home |

\*Status unavailable at the time of reportin

**Table 3:** Outcome of COVID-19 Patients Treated with Complete 7 Days Course DOXY-HCQ Combination in Three (3) Long Term Care Facilities^*^.

| Characteristics | LTCF Resident (n=45) | % | Reference Population (LTCF,WA) |
| --- | --- | --- | --- |
| Median Age, yrs. (range) | 68 (22-97) |  |  |
| **Chronic underlying conditions** |  |  |  |
| Hypertension | 41 | 91% |  |
| Diabetes | 18 | 40% |  |
| CAD | 26 | 58% |  |
| CHF | 8 | 18% |  |
| CVA | 17 | 38% |  |
| COPD | 17 | 38% |  |
| Vent dependent | 12 | 27% |  |
| **Outcome** |  | 0% |  |
| Clinically recovered** | 45 | 100% |  |
| Transferred to hospital | 0 | 0% | 57% |
| Death | 0 | 0% | 22% |
| **Radiology** |  | 0% |  |
| Abnormal CXR c/w Pneumonia | 28 | 62% |  |
| Chest x-ray improved | 5 | 11% |  |
| Chest x-ray not improved (possible lag time) | 10 | 22% |  |
| Unavailable*** | 30 | 67% |  |
| **Medication side-effect** |  | 0% |  |
| None | 45 | 100% |  |

\* This table includes all patients who completed a full course (7 day) of DOXY-HCQ therapy and excludes any patients who were transferred to hospital, death or developed side-effects due to DOXY-HCQ and did not complete the 7-day course.
\*\* Recovery defined by resolution of fever, shortness of breath or return to baseline vent setting
\*\*\* Chest x-ray either not ordered, or not required or pending

## Discussion

The COVID-19 pandemic created a huge burden on the U.S. healthcare system and increased the risk of hospitalization, ICU admissions, and mortality. Recently, various drugs have been experimentally used, and the most promising therapy involves HCQ and/or AZ. However, safety concerns are raised due to the potential side-effects profile of both drugs, particularly when used in combination.

A recent open-label French study of only six (6) patients in hospital settings showed limited effectiveness (no clinical outcomes reported) of combination of AZ-HCQ in treatment of COVID-19 patients, within six days of treatment^8^. Subsequently, Raoult et al^17^ published another case series of 80 patients in hospital settings with AZ-HCQ combination therapy and all but two patients showed clinical improvement. However, 94% of patients were low-severity patients as evidenced by low (0-4) national early warning score (NEWS). The impact of such combination therapy in a high-risk population like ours is unknown.

Another recently published Chinese randomized clinical trial conducted in a hospital setting among 62 patients treated with HCQ for 5 days showed improvement of pneumonia in the HCQ treatment group (80.6%, n=25 of 31 patients) compared with the control group (54.8%, n=17 of 31 patients)^6^. 2 patients showed mild adverse events including rash and headache. Detailed demographics of the patient population were not published, however the mean (SD) age was 44.7 (15.3) yrs., much younger than our patients. Compared to this study, our case series patients were much older and had significant multiple comorbidities.

So far, no study was published from the United States using HCQ alone or in combination with AZ or DOXY. As a matter of fact, this study is the first case series conducted globally using a combination of DOXY-HCQ in COVID-19 patients in nursing home settings. We treated our moderate to severe patients in out-of-hospital settings and prevented hospital transfer in the majority of patients. Naive Indirect Comparison showed a better outcome than the data reported in MMWR (reported on March 26, 2020)^17^ from a long-term care facility in King County, Washington where 57% of patients were hospitalized and 22% patients died. This effect can possibly be explained by the anti-inflammatory mechanism of action of DOXY-HCQ.

However, data from the LTCF from King County, Washington was found to be similar to our population. These data also showed a reduction of hospitalization by 44% among LTCF residents compared to previously reported data by similar populations in MMWR.

A limitation of our case-series analysis is that there was no control population available.

## Conclusion

DOXY and HCQ combination therapy is known to be anti-inflammatory, and immunomodulatory in both in-vitro and in-vivo studies. In addition, HCQ has anti-viral properties. Although this sample size is small (n=54), the results suggest that early intervention of DOXY-HCQ may improve the clinical outcome of high-risk COVID-19 patients suffering from moderate-severe symptoms in LTCF. These data is also associated with a reduction of hospitalization by 44% among moderate to high severity COVID-19 LTCF residents compared with previously reported data by similar populations from King county, Washington^18^.

Given the massive pandemic created by COVID-19 virus, there is an emergent therapeutic need to manage this disease with effective and safe drugs in an out-of-hospital setting for high-risk patients to reduce hospitalization and ICU admission and manage acute ventilator shortage. A well-designed clinical study is urgently needed to identify the appropriate patient population, optimize dosing regimen, and assess side-effects of DOXY-HCQ combination therapy.

## Data Availability

All data available from corresponding author

